# Exploring Early Perceptions and Experiences of ChatGPT in Pediatric Critical Care: A Qualitative Study Among Healthcare Professionals

**DOI:** 10.1101/2024.03.18.24304453

**Authors:** Mohamad-Hani Temsah, Noura Abouammoh, Mohammed Alsatrawi, Mohammed Almazyad, Fadi Aljamaan, Mariella Vargas-Gutierrez, Rebecca Hay, Muneera Al-Jelaify, Wejdan Alabdulkreem, Nawal Assiri, Ruaim Muaygil, Ibraheem Altamimi, Fatimah S. Alshahrani, Khalid Alhasan, Khalid H. Malki, Amr Jamal, Jaffar A. Al-Tawfiq, Ayman Al-Eyadhy

**Author notes:** Corresponding author: Mohamad-Hani Temsah. **Author Contributions:** Mohamad-Hani Temsah, Noura Abouammoh, Mohammed Almazyad, Mariella Vargas-Gutierrez, Rebecca Hay and Mohammed Alsatrawi roles were conceptualization, data curation, formal analysis, funding acquisition, investigation, methodology, project administration, resources, software, supervision, validation, visualization, writing – original draft, and writing – review & editing the final version. All authors directly accessed and verified the underlying data reported in the manuscript. Muneera Al-Jelaify, Wejdan Alabdulkareem, Nawal Assiri, Fadi Aljamaan, Khalid Alhasan, Khalid H. Malki, Ibraheem Altamimi, Fatimah Alshahrani, Ruaim Muaygil, Amr Jamal, Jaffar A. Al-Tawfiq and Ayman Al-Eyadhy contributed to the data curation, investigation, methodology, resources, software, validation, visualization, writing – original draft, and writing – review & editing the final version. All authors have read and agreed to the published version of the manuscript. **Funding:** This research received no external funding. **Data Availability Statement:** The deidentified participant data collected for this study will be made available to others, upon reasonable request, from the corresponding author, after approval of a proposal, in agreement with the IRB-provided signed data sharing agreement.

## Abstract

This qualitative inquiry explores the initial impressions and firsthand encounters of healthcare professionals (HCPs) with ChatGPT, a Generative Pre-trained Transformer, within Pediatric Intensive Care Units (PICUs). Through focus group discussions held at a tertiary academic center, a diverse cadre of HCPs was engaged to ascertain their awareness, utilization patterns, perceived advantages, and apprehensions regarding ChatGPT. The analysis revealed three primary themes: understanding and ease of use of ChatGPT, its practical applications in clinical workflows for critically ill children and information retrieval, and the ethical considerations associated with its deployment. While participants praised ChatGPT for its engaging interface and potential to streamline tasks and provide prompt information, notable reservations surfaced regarding its limitations, particularly in medical accuracy, currency of data, and ethical implications. The findings suggest a cautious optimism towards integrating Generative Artificial Intelligence (GAI), like ChatGPT, in pediatric critical care, highlighting the need for balanced, informed, and transparent applications, with ongoing evaluation of GAI technologies in pediatric healthcare settings.

## Introduction

Artificial intelligence (AI) applications are increasingly prevalent in society, becoming integral to our daily lives. In medicine, AI is poised to significantly transform medical practice and patient care. Furthermore, the generative AI language models, or Large Language Models (LLMs), are becoming widely available starting in early 2023[1]. Integration of such LLM has shown promise in facilitating discussions in pediatric palliative care panels [2]. ChatGPT, the Generative Pre-trained Transformer (GPT), created by OpenAI, is a model for generating natural language that predicts subsequent words in text sequences to produce content closely mirroring human writing. ChatGPT was launched in Nov 2022 and garnered over 100 million users within two months[3]. GPT-3, with its 175 billion parameters—a tenfold increase over any prior dense language model— GPT-3, boasting 175 billion parameters— representing a tenfold expansion compared to any previous dense language model—is an autoregressive model that has seen widespread adoption and rapid integration into public applications, leading to its frequent discussion and publication in diverse medical literature [4,5].

In March 2023, the enhanced version, GPT-4, was released, featuring significant improvements in efficiency [6]. Newer models introduced further potentials to be used in formulating management plans for diverse patient populations and scenarios, with pros and cons extensively published in ever expanding medical literature [5]. Still, the growing role of AI in medicine is not without controversy and misunderstanding [7-9]. Concerns include the potential erosion of doctors’ skills over time, impacts on the doctor-patient relationship, and issues related to data privacy, patient autonomy, and informed consent [10,11].

Recent studies have sought to explore healthcare professionals’ attitudes and views towards generative AI and its applications in their medical practice. Parikh et al. conducted an online cross-sectional descriptive survey in Feb 2023, revealed that less than 50% of healthcare professionals had utilized ChatGPT [12]. While they generally held a positive view of it, a significant majority (80%) did not anticipate it would have a substantial impact on their practice at least in 2023 (8). Another study in Saudi Arabia found that most healthcare workers believed ChatGPT could positively affect the future of healthcare systems; however, serious concerns were raised about its credibility [13].

Healthcare workers (HCWs) in PICUs face constant challenges in managing diverse and evolving critical illnesses in vulnerable pediatric age group, striving to make timely clinical decisions and interventions aiming to improve morbidity and mortality. Communications in PICU are always challenging, whether in-between medical teams or between HCWs and families [14]. Recently, generative AI models have been employed in multiple domains of intensive care, especially in relation to diagnosis and prognosis prediction and management, interestingly, even assisting in goals of care measurement by summarizing and processing clinical notes and data [15-21]. Another valuable use of AI model is teaching, training and skills development in safe environment as circulatory shock detection and management, using simulation technique facilitated by AI [22]. Therefore, generative AI models like ChatGPT might be a potential tool assisting practitioners in PICU in various aspects, such as foreseeing complications, complex decision-making by deciphering intricate patterns of complex data sets, on the other hand facilitating communication process at multiple levels.

This qualitative study aims to evaluate healthcare professionals in the PICU setting, in terms of their awareness and perceptions of generative AI technologies, particularly ChatGPT, as it was not probed yet in literature to the best of our knowledge, offering specific insights that can guide the creation, application, and enhancement of AI chatbots in pediatric critical care. Comprehensively understanding ChatGPT’s potential benefits and limitations within specific field of healthcare practice would offer insights facilitating more effective and safer integration of similar LLMs into pediatric critical care ecosystems, addressing more focused comprehension and prediction of future perspectives concerning possible advantages and constraints. These observations can aid in the development of guidelines and the making of educated decisions by developers, legislators, and healthcare professionals regarding the successful adoption and application of generative AI models in pediatric critical care.

## Methods

This qualitative study was conducted at King Saud University Medical City, Riyadh, Saudi Arabia, a tertiary academic center with two PICUs at King Khalid University Hospital and King Abdulaziz University Hospital, Riyadh, Saudi Arabia. Participants were selected through purposive sampling from these two PICUs locations. The focus group discussions included healthcare professionals from various backgrounds working with critically ill children, including physicians with various roles, nurses, pharmacists, dieticians, social services, and a hospital ethicist.

The sample comprised individuals with diverse experiences with ChatGPT, ranging from scarce uses to regular usage. Two focus group discussions were held in May 2023 and August 2023, via Zoom platform, each group having nine participants. The focus group discussions were conducted in English, as it is the official communication language within the hospital, moderated by two authors (NA, MHT), and each lasted approximately one hour. To ensure precision, the discussions were audio-recorded and transcribed verbatim.

Data was collected using semi-structured focus group discussions. This technique in data collection was used as it allows the researcher to explore range of ideas and perspectives and easily reveals differences and similarities between participants and factors influencing participants’ attitude through moderated group interaction [23,24]. Focus group discussion is also appropriate in exploring topics where minimal literature is known, which is applicable in this study as attitudes and uses of ChatGPT in clinical settings is still unclear [24,25]. Virtual technique was used to allow participant recruitment giving their busy schedule, offering possible advantages for participants’ diversity and outreach [25-27].

The discussions were conducted using a topic guide. Questions included participants’ familiarity with the ChatGPT, its uses, and the facilitators and barriers to its integration in medical education. Probing and follow-up questions were tailored to participants’ responses. Theme saturation was achieved after the second interview. Thematic analysis was employed to examine the data, using both priori themes and allowing new themes to emerge. Transcripts were repeatedly reviewed to identify patterns and themes. A coding framework was applied using NVivo 12 software [28]. Themes were identified and refined through a continuous process of coding, reviewing, and discussing the data within the research team until a consensus and themes saturation were reached.

Ethical approval was granted by the Institutional Review Board at the participating university (Ref. No. 23/0155/IRB). Verbal informed consent was obtained from all participants regarding their participation and the audio recording of the discussions. They were informed about the study’s purpose, the voluntary nature of their involvement, and their right to withdraw at any time. Pseudonyms were employed to maintain participant anonymity.

## Results

Table 1 provides an overview of the demographic details of the participants, reflecting a diverse representation across various roles within the pediatric critical care setting. The participants included PICU and pediatric consultants, PICU fellows, clinical pharmacists, PICU nurses, a social worker, an ethicist, and a dietitian. This broad spectrum of roles aimed to have a comprehensive exploration of early perceptions and experiences regarding the utilization of ChatGPT in pediatric critical care. Three primary themes emerged from the focus group discussions with the PICU team (Table 2 and Fig 1): knowledge about ChatGPT among the PICU team, uses of ChatGPT in the PICU, and ethical concerns regarding ChatGPT usage.

**Table 1:** Demographic information of the focus group participants.

| PARTICIPANT | PICU TEAM ROLES | GENDER | CHATGPT USE |
| --- | --- | --- | --- |
| P1 | Consultant PICU | Male | Yes: Weekly |
| P2 | Consultant ethicist | Female | No |
| P3 | Consultant PICU | Male | Yes: Daily |
| P4 | Clinical Pharmacist | Female | Yes: Weekly |
| P5 | Pediatric Consultant | Female | Yes: Weekly |
| P6 | PICU Fellow | Male | Yes: Weekly |
| P7 | Consultant PICU | Male | Yes: Occasional |
| P8 | PICU Senior | Female | Yes: Daily |
| P9 | Clinical Pharmacist | Female | Yes: Occasional |
| P10 | Dietitian | Female | Yes: Weekly |
| P11 | Clinical Pharmacist | Female | Yes: Daily |
| P12 | PICU Fellow | Female | Yes: Daily |
| P13 | PICU Fellow | Female | Yes: Occasional |
| P14 | PICU Head Nurse | Female | Yes: Occasional |
| P15 | PICU Charge Nurse | Female | No |
| P16 | PICU Social Worker | Female | Yes: Occasional |
| P17 | PICU Consultants | Male | Yes: Occasional |
| P18 | Pediatric Consultation service | Male | Yes: weekly |

**Table 2:** Process of deriving themes from the focus group discussion.

| Primary code | Category | Theme |
| --- | --- | --- |
| User friendly<br>Limitations of chat GPT | Participants acknowledge that ChatGPT is user friendly<br>Limitations of ChatGPT included evolution process, sources of the information | Knowledge about ChatGPT among PICU team |
|  | displayed and recency of data |  |
| • ChatGPT as an organizer<br>• ChatGPT as a source of information in practice | 1. ChatGPT is reliable in organizing tasks<br>2. ChatGPT is reliable and quick to gain general information<br>3. ChatGPT confirm information participants' already knew<br>4. Participants would not recommend ChatGPT use to obtain medical information among patients and caregivers | Uses of ChatGPT among PICU team |
| • ChatGPT and ethics | Privacy concerns<br>Communication concerns | Ethical Concerns When Using ChatGPT |

**Figure 1:**
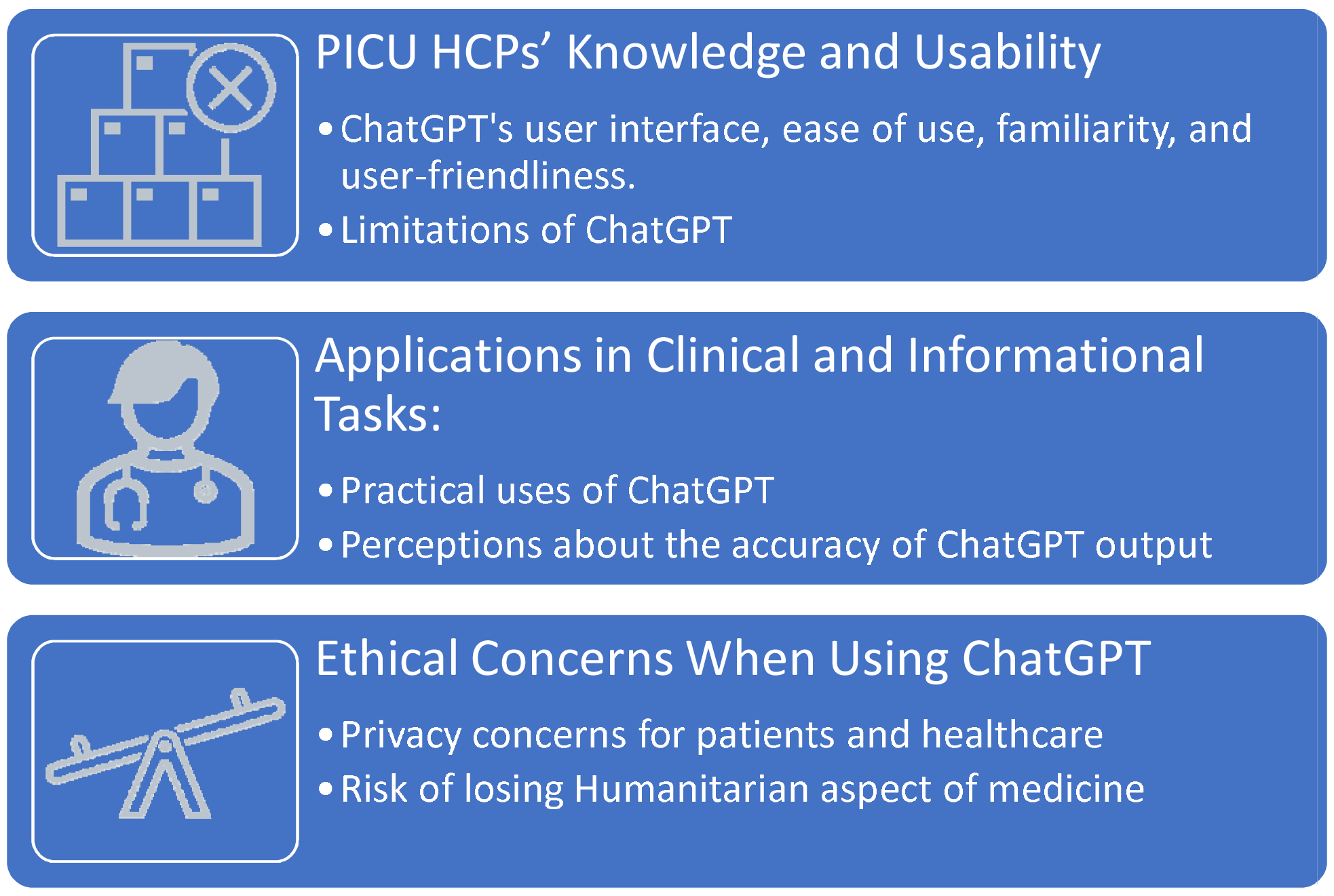
Perceptions and Applications of ChatGPT in Pediatric Intensive Care

## 1 Knowledge about ChatGPT among PICU team

In our focused-group discussion of the PICU team’s knowledge about ChatGPT, it became evident that most participants were familiar with its functionalities and found it to be highly user-friendly. Their experiences echoed an ease of use akin to conversing with a person, as detailed below. However, participants also recognized limitations, notably its reliance on pre-training datasets and the potential for outdated information due to its last training cycle or unknown database sources. These insights highlight both the strengths and challenges associated with integrating ChatGPT into pediatric critical care contexts.

### 1.1 User friendly

All participants (16/18) who used ChatGPT acknowledged its user-friendliness and were familiar with its functionalities and human-like interactions. For instance, one participant commented, “This technology is easy to use. It can answer questions as if I am talking to a person.” (P6)

Another participant clarified, “It is not a search engine. It doesn’t generate new information but predicts the most probable or appropriate word in the given context.” (P3)

Additionally, a participant described it as a processor, stating, “It processes the information it has access to and presents it in a well-written language format, adding a conversational dimension to ChatGPT.” (P10)

One healthcare worker (HCW) favorably compared ChatGPT to Google, noting, “You can speak to it as if you’re discussing a topic with someone… and it supports multiple languages.” (P14)

### 1.2 Limitations of ChatGPT

Participants recognized one current limitation of ChatGPT; describing it as a ‘limited’ source due to its pre-training on specific datasets, and its ongoing evolution based on user’s interactions and prompting. One participant pointed out, “If ChatGPT receives incorrect information, it can update itself and seek new resources and information.” (P6)

Participants also noted limitations related to the recency of data, referring to the ChatGPT-3.5 being trained on data until Sept 2021 at the time of the interview [29]. One participant observed, “The data is limited to information from up to two years ago, so it might not provide the most recent updates.” (P7)

## 2 Uses of ChatGPT among PICU team

In exploring the uses of ChatGPT among the PICU team, participants identified its role as both a capable organizer and an effective source of information. They praised its ability to streamline tasks and provide quick responses to inquiries during clinical rounds. However, concerns were raised regarding its accuracy and reliability in medical contexts, highlighting the need for cautious evaluation and vigilance when applying its capabilities in clinical practice.

### 2.1 ChatGPT as an Organizer

ChatGPT was frequently mentioned as a tool for organizing tasks. One participant expressed it as, “It’s like security.” (P9) Another highlighted ChatGPT’s role in daily planning: “I write my to-do list and then ask ChatGPT to propose a schedule for me to finish the tasks during the day.” (P17) The same participant also mentioned using ChatGPT for weekly planning, stating, “I tell ChatGPT about my patient rounds, weekly meetings, and research office hours, and it suggests a comprehensive schedule, including breaks and lunchtime.” (P17)

### 2.2 ChatGPT as a Source of Information in Practice

#### 2.2.1 Practical uses of ChatGPT

All participants agreed that ChatGPT serves as a “reliable” source of information for general inquiries and appreciated its quick responses. For example, a participant mentioned, “Sometimes I use ChatGPT during rounds if I need a quick reminder.” (P11)

A pharmacist added, “It provides me with drug dosages as this information is fairly standardized.” (P7)

ChatGPT was also utilized for brainstorming. A consultant stated, “It’s a great tool for generating ideas, whether for quality assessments or research projects. It offers numerous suggestions, which help in opening up new perspectives.” (P7)

Furthermore, a senior PICU member mentioned using ChatGPT to supplement her knowledge: “I presented a clinical question about a real-life patient scenario and provisional diagnosis. ChatGPT suggested a list of appropriate actions for the case. However, due to the variable nature of ICU patient management, solid background knowledge is essential to determine the best course of action.” (P8)

Participants also viewed ChatGPT as a helpful guide for initiating research. For instance, one participant said, “Sometimes, I am unsure where to start a search, especially if Google does not yield results. ChatGPT provides a starting point.” (P11)

It was observed that participants generally used ChatGPT to confirm information they already knew or answering medical questions during the PICU clinical rounds: “It is useful for quick reference or to double-check information for concise answers,” noted a participant (P11).

However, one participant articulated a clear perspective on ChatGPT’s appropriate use, a view supported by others in both focus groups: “ChatGPT is not designed for acquiring knowledge, understanding subjects, or aiding physicians in decision-making, limiting its utility for these purposes. It’s beneficial for writing recommendations or articles and analyzing some data.” (P3)

This participant also cautioned, “One might be swayed by how ChatGPT presents information – usually in eloquent language.” (P3) This highlights an important consideration regarding the perceived accuracy and reliability of the information provided by ChatGPT.

### 2.2.2 Perception about Caregivers’ Use of ChatGPT

Most participants expressed skepticism about recommending ChatGPT as an information source for families and caregivers. One participant cautioned, “It might mislead them more than guide them, and ChatGPT performs worse in Arabic than in English. I’d suggest they consult other sources.” (P7)

Another added, “I wouldn’t recommend ChatGPT to my patients. Given my own reservations about its medical accuracy, I can’t imagine adding ChatGPT-sourced information to the mix of questions patients already bring from Google searches.” (P9)

The social worker in the group had tested ChatGPT in Arabic to gauge its utility for patients and caregivers: “The responses in Arabic are too general. It offers an overview rather than detailed information, so I wouldn’t recommend it for families.” (P16)

Conversely, a physician shared a positive experience: “A relative used ChatGPT to interpret an MRI report, and its response was similar to mine. Importantly, it advised consulting a doctor.” (P13)

A PICU consultant saw no harm in families using ChatGPT for health information, provided there is trust in the medical team: “Patients will seek information if it is accessible. They return to us for verification. It differs from those who have lost trust and rely solely on the web.” (P3)

To address concerns, this consultant suggested, “We should offer educational materials on common diseases we encounter to patients as alternatives.” (P3)

Overall, participants acknowledged that the Saudi community uses ChatGPT, though not predominantly for medical information.

### 2.2.3 Perceptions about the accuracy of ChatGPT output

Participants shared experiences with ChatGPT that raised concerns about the accuracy and reliability of its output, particularly in medical contexts. They noted issues with both the precision of information and the language used in responses.

One participant recounted an instance where ChatGPT provided a correct equation for creatinine clearance but yielded an incorrect result: “I tried a simple calculation… ChatGPT gave me the right equation but the wrong answer.” (P11)

Another highlighted the need for expertise in evaluating ChatGPT’s outputs, especially for medication dosages: “Seniority is required to decide whether to rely on ChatGPT’s calculations. I would use it for understanding medication mechanisms and side effects.” (P12)

A PICU fellow observed that ChatGPT’s usefulness of its responses vary based on how questions are posed: “If I specify that I’m a physician asking about a drug’s mechanism, the response is more informative than a general query.” (P13)

Participants also expressed concerns about ChatGPT’s handling of controversial health topics: “Sometimes there’s controversy in practice, and ChatGPT doesn’t address it well.” (P7)

The issue of providing reliable references was another concern: “ChatGPT sometimes cites fake links or provides inaccurate references not found in the literature.” (P7, P9)

While one participant noted ChatGPT’s ability to provide references, the overall sentiment was skepticism due to its conversational style: “Information is presented conversationally, unlike typical medical data formats, which triggers doubts about its reliability.” (P10)

This skepticism extended to relying on ChatGPT for clinical decision-making: “I wouldn’t jeopardize patient health by relying on ChatGPT. I’d prefer traditional, reliable information sources.” (P7)

The fact that ChatGPT often concludes with a suggestion to consult an expert further challenges its perceived reliability.

Overall, participants tested ChatGPT with known information to gauge its efficiency and determine its utility in their practice.

## 3. Ethical Concerns When Using ChatGPT

Participants in our study raised ethical concerns regarding the integration of ChatGPT in healthcare practice. While some emphasized the need for caution due to the technology’s novelty and potential impact on patient empathy and privacy, others argued for its exploration to keep pace with advancing healthcare technologies. Concerns about patient communication dynamics and data privacy were prominent, prompting calls for balanced regulation to ensure ChatGPT complements rather than supplants human expertise while upholding ethical standards in healthcare delivery.

A clinical ethicist among the participants expressed strong reservations about prematurely employing ChatGPT in healthcare, citing its novelty, potential privacy issues, and the risk of losing compassion and empathy in medical practice: “We shouldn’t hastily embrace new technologies without considering their moral implications. The unknowns about ChatGPT outnumber the knowns.” (P2)

Another participant responded to this cautious stance, emphasizing the need to adapt: “Exploring new technologies like ChatGPT is essential. Otherwise, we risk falling behind in the coming years.” (P10)

Concerns were also expressed about ChatGPT affecting crucial aspects of patient communication, such as sympathy and empathy: “Incorporating ChatGPT might erode the humanitarian aspect of medicine.” (P2)

The potential breach of patient privacy was a significant worry. While opinions varied, one participant emphasized the risks associated with AI and data privacy: “Reliance on AI, like ChatGPT, could compromise personal privacy and agency, potentially leading to the creation of databases with patient information, even if identities are obscured.” (P2)

Another participant, agreeing with the critique of AI but sensing resistance, suggested a more balanced approach: “AI is here to stay. We shouldn’t resist it but rather regulate it. It should be seen as an aid, not a replacement in human medicine.” (P3)

These discussions reflect a nuanced understanding of ChatGPT’s role in healthcare, recognizing its potential benefits while being acutely aware of the ethical implications and the need for careful regulation and oversight.

## Discussion

Rapid advancements in AI technologies are transforming medical practices across various medical fields. AI has the capability to help diagnose illnesses, create individualized treatment strategies, and support healthcare professionals in decision-making processes [30,31]. Its application goes beyond mere task automation, aiming to advance patient care throughout various healthcare environments. . In another domain, AI models were developed and tested to predict potential COVID-19 infections and prognosis with high accuracy [32,33]. The application of AI is also continuously expanding into pediatric field and PICUs. Dong et al. developed a model for predicting acute kidney injury (AKI) in pediatric critical care, capable of accurate predictions up to 48 hours before AKI onset [34]. The recent launch of publicly available generative AI tools, like ChatGPT, would have additional impacts in healthcare, as this becomes a more integral part of HCWs and patients’ dynamics.

The results of our focus group discussions delineate a multifaceted view of ChatGPT among PICU healthcare professionals. The findings underscore a dichotomy in perspectives: while recognizing the potential utility of ChatGPT in streamlining tasks and providing informational support, participants expressed significant reservations about its limitations, particularly regarding data recency, accuracy, and the potential impact on the healthcare professional-patient relationship. The consensus on ChatGPT’s user-friendliness and its conversational, multi-lingual capabilities illustrate an appreciation of the technology’s ergonomic design and accessibility. This aligns with the growing trend of AI integration in healthcare, where ease of use is paramount for widespread adoption [35]. However, the participants’ perception of ChatGPT as a “limited” source, primarily due to its dependence on pre-existing datasets and the absence of real-time and accurate data updates, highlights a critical aspect of AI in healthcare: the need for continuous AI models in learning and adaptation to verified yet evolving medical knowledge [36].

The application of ChatGPT as an organizer and information source reflects its perceived role as an adjunct, rather than a replacement, in clinical decision-making processes [36]. This is particularly noteworthy in the context of pediatric intensive care, where the complexity and variability of cases necessitate human expertise and judgment. The participants’ reliance on ChatGPT for confirming known information rather than as a primary source underscores the need for AI tools to be used as supplements to, rather than substitutes for, human clinical judgment. The skepticism about recommending ChatGPT for use by caregivers and patients, primarily due to concerns about medical accuracy and linguistic limitations, highlights the need for caution in the deployment of AI tools in patient education and communication. This finding echoes concerns raised in other studies about the potential for misinformation and misunderstanding when AI tools are used without proper context or guidance [37].

In pediatric critical care, AI’s role has become pivotal in overcoming the limitations posed by heterogeneity across age groups and the challenges in defining and subclassifying disease phenotypes. Machine learning algorithms assist in generating meaningful interpretations from clinical data, thereby enhancing clinical decision-making at the bedside. This is especially crucial given the large amounts of underutilized data generated in critical care units. AI’s role extends beyond mere data analysis to include offering evidence-based recommendations for critically ill pediatric patients and improving the predictive performance of underlying models, especially for high-dimensional data [38,39].

In this study, participants raised several ethical concerns regarding the use of ChatGPT. The deployment of AI in healthcare, including ChatGPT, raises significant ethical considerations. The distinction between AI as electronic expert systems used by healthcare professionals and full AIs, which function independently, highlights the ethical ambiguity surrounding AI’s application in healthcare. These considerations include issues of data privacy, patient autonomy, and the potential for AI to erode the human elements of healthcare. The ethical implications of AI necessitate robust regulatory measures to ensure that its deployment in healthcare sectors is balanced, preserving compassionate and empathetic patient care. This balance is crucial to maintaining the integrity of healthcare practices and safeguarding patient interests [40].

The ethical concerns raised by the participants, particularly regarding patient privacy and the potential erosion of the humanitarian aspects of medicine, highlight the need for a balanced approach to AI integration in healthcare. This aligns with the broader discourse on AI ethics, which emphasizes the importance of developing AI technologies in a way that respects patient autonomy, confidentiality, and the humanistic values of medicine. The participants’ varied responses to the potential of ChatGPT in healthcare reflect a broader uncertainty within the medical community about the role of AI. This underscores the importance of continued dialogue and research to explore the implications of AI in healthcare, particularly in sensitive areas such as pediatric intensive care [41].

ChatGPT’s performance in medical exams and its implications in medical education also relate to the academic functioning in the PICU setting [3]. A study by Gilson et al., evaluated ChatGPT’s performance on the United States Medical Licensing Examination, revealing its potential as an interactive tool for medical education [42]. Their study demonstrated that ChatGPT could perform at a level equivalent to a passing score for a third-year medical student, suggesting its potential utility in critical care for educational purposes. However, the study also highlighted the model’s limitations, especially in handling more complex, high-difficulty questions, underscoring the need for continuous evolution and oversight in its application in medical education and training [42].

Also, communication is vital among HCWs, patients and families critical care but is frequently inadequate in the PICU setting [43]. Communication training increases the PICU fellows’ confidence in having difficult discussions, and LLMs could be used to facilitate such skills, like training AI models as interactive simulation in breaking bad news to families [44,45]. Future AI chatbot models could also improve the clinical reasoning of HCWs, though more research is warranted [46].

A systematic review outlined both the benefits and risks associated with ChatGPT’s use in healthcare [47]. The review highlighted ChatGPT’s utility in scientific writing, efficient analysis of massive datasets, and support in drug discovery and development. However, it also emphasized significant concerns regarding ethical issues, risk of bias, and potential for incorrect responses. In the PICU context, while ChatGPT can streamline workflow and support research, these limitations necessitate cautious and informed application to ensure patient safety and data integrity.

The ability of ChatGPT to provide appropriate and equitable medical advice was evaluated by Nastasi et al., as they presented ChatGPT with various clinical vignettes and found that while ChatGPT’s responses were largely in line with clinical guidelines, it did not consistently offer personalized medical advice [48]. AI algorithms carry inherent dangers that may lead to incorrect conclusions and operate as “black box” systems that do not elucidate the rationale behind decision-making [49]. Saenger et al. present a case report of a patient who relied on ChatGPT inaccurate diagnosis, and that resulted in a substantial delay in treatment and a potentially life-threatening circumstance [49]. These findings raise important clinical and ethical considerations, especially in the sensitive setting of a PICU, where decisions can have significant consequences. It emphasizes the need for human oversight and the importance of not overly relying on AI tools like ChatGPT for critical healthcare decisions. Trust in ChatGPT’s medical advice, especially in complex pediatric cases, must be balanced with an understanding of its limitations and the necessity of human expertise in clinical judgment [48,50-52].

## Strengths and Limitations

The study, while insightful during the initial months of ChatGPT usage among HCWs, is subject to certain limitations. Firstly, the small sample size may restrict the generalizability of the findings across different PICU settings. Additionally, the study did not employ quantitative methods to evaluate ChatGPT’s impact on patient outcomes, satisfaction, or other quantifiable aspects of PICU operations. Future research with such data would offer additional insights to complement the qualitative findings.

Another limitation is the study’s focus solely on the perspectives of HCWs, excluding views from other important stakeholders like computer scientists and policymakers. This exclusion could impact the completeness of the study’s findings and warrants further comprehensive strategies for researching optimal integrating ChatGPT into critical care and hospital settings.

Moreover, the study did not investigate the long-term effects of ChatGPT usage and its evolving perceptions among users. As experiences with the tool and newer LLMs models change over time, so might the assessments of its advantages and drawbacks, and ongoing assessments are needed.

Future research should aim to mitigate these limitations. Expanding the sample size and including diverse participants from various PICUs would enhance the representativeness of the findings. Quantitative studies are also essential to objectively measure the impact of ChatGPT on different facets of medical practice. Including the perspectives of other stakeholders, such as administrators and policymakers, would provide a more holistic understanding of the implications of integrating ChatGPT. Finally, longitudinal studies could track the evolving perceptions and uses of ChatGPT over time, shedding light on its long-term effects in medical education and practice.

## Conclusion

Our qualitative study offers important insights into how pediatric critical care healthcare professionals view and use ChatGPT, a popular artificial intelligence (AI) chatbot. It presents a nuanced picture in which ChatGPT is acknowledged for its ability to streamline processes, such as clinical decision making, and provide informational assistance, but it also raises serious questions regarding its limitations in terms of data recency, accuracy, ethics, and impact on the patient-physician relationship. The findings highlight the growing significance of AI in healthcare, but they additionally emphasize the necessity to maintain the humanistic elements of patient care while balancing technological innovation with ethical considerations. Our study stresses the necessity of continued investigation and a cautious, multidisciplinary approach when incorporating ChatGPT and other AI instruments into sensitive and intricate medical domains.

## Data Availability

All data produced in the present study are available upon reasonable request to the corresponding author

## Acknowledgments

We have used ChatGPT, an AI chatbot developed by OpenAI, to improve the readability and language of this work without replacing researchers’ tasks. This was done with human oversight, and authors then carefully reviewed and edited the generated text, as we assure that the authors are ultimately responsible and accountable for the originality, accuracy, and integrity of their work. We would like to acknowledge the efforts in data curation in the focus groups. The authors extend their appreciation to the Deputyship for Research and Innovation, Ministry of Education, Saudi Arabia, for supporting this research (IFKSURC-1-3110).

